# Interest of tri-axial rubidium and helium optically pumped magnetometers for on-scalp magnetoencephalography recording of interictal epileptiform discharges

**DOI:** 10.1101/2023.10.02.23296348

**Authors:** Odile Feys, Pierre Corvilain, Etienne Labyt, Mahdi Mahmoudzadeh, Laura Routier, Claudine Sculier, Niall Holmes, Matthew Brookes, Serge Goldman, Rudy Romain, Sergey Mitryukovskiy, Agustin Palacios-Laloy, Denis Schwartz, Nacim Betrouni, Philippe Derambure, Fabrice Wallois, Vincent Wens, Xavier De Tiège

**Affiliations:** Université libre de Bruxelles (ULB), Hôpital Universitaire de Bruxelles (HUB), CUB Hôpital Erasme, Department of Neurology, Brussels, Belgium; Université libre de Bruxelles (ULB), ULB Neuroscience Institute (UNI), Laboratoire de Neuroimagerie et Neuroanatomie translationnelles (LN^2^T), Brussels, Belgium; Mag4Health, Grenoble, France; CEA Tech en Hauts de France, EuraTechnologies, Lille, France; Centre Hospitalier Universitaire Amiens-Picardie, Inserm, U1105, GRAMFC, Explorations Fonctionnelles du Système Nerveux Pédiatrique, Amiens, France; Université libre de Bruxelles (ULB), Hôpital Universitaire de Bruxelles (HUB), CUB Hôpital Erasme, Department of Pediatric Neurology, Brussels, Belgium; University of Nottingham, School of Physics and Astronomy, Sir Peter Mansfield Imaging Centre, Nottingham, United Kingdom; Université libre de Bruxelles (ULB), Hôpital Universitaire de Bruxelles (HUB), CUB Hôpital Erasme, Department of Nuclear Medicine, Brussels, Belgium; CERMEP-Imagerie du Vivant, MEG Departement, Lyon, France; Université de Lille, INSERM, U1172 – LilNCog – Lille Neuroscience & Cognition, F-59000, Lille, France; Université libre de Bruxelles (ULB), Hôpital Universitaire de Bruxelles (HUB), CUB Hôpital Erasme, Department of Translational Neuroimaging, Brussels, Belgium

**Keywords:** Magnetoencephalography, Optically pumped magnetometers, On-scalp magnetoencephalography, Refractory epilepsy, Focal epilepsy

## Abstract

Cryogenic magnetoencephalography (MEG) enhances the presurgical assessment of refractory focal epilepsy (RFE). Optically pumped magnetometers (OPMs) are cryogen-free sensors that enable on-scalp MEG recordings. Here, we investigate the interest of tri-axial OPMs (^87^Rb (Rb-OPM) and ^4^He gas (He-OPM)) for the detection of interictal epileptiform discharges (IEDs).

IEDs were recorded simultaneously with 4 tri-axial Rb- and 4 tri-axial He-OPMs in a child with RFE. IEDs were identified visually, isolated from magnetic background noise using independent component analysis (ICA), and the orientation of magnetic field generated by the IEDs was reconstructed at each sensor location.

Most IEDs (>1,000) were detectable by both He- and Rb-OPM recordings. IEDs were isolated by ICA and the resulting magnetic field oriented mostly tangential to the scalp in Rb-OPMs and radial in He-OPMs. Likely due to differences in sensor locations, the IED amplitude was higher with Rb-OPMs.

This case study shows comparable ability of Rb-OPMs and He-OPMs to detect IEDs and the substantial benefits of triaxial OPMs to detect IEDs from different sensor locations. Tri-axial OPMs allow to maximize spatial brain sampling for IEDs detection with a limited number of sensors.

## Introduction

Magnetoencephalography (MEG) records the magnetic fields generated by electrical brain activity (1). Its main clinical application is the non-invasive presurgical evaluation of refractory focal epilepsy (RFE) (2).

Cryogenic MEG systems are typically based on ∼300 superconducting quantum interference devices (SQUIDs) (3) requiring cryogenic cooling in liquid Helium (−269°C) to record neuromagnetic fields (4). Sensors are thus housed in a one-size-fits-all, commonly adult-sized, helmet to maintain a thermal isolation space (2–3 cm) with the scalp (3), therefore reducing the signal-to-noise ratio (SNR) (5). This issue is *a fortiori* even worse for subjects with a small head circumference such as children (3).

Optically pumped magnetometers (OPMs) are novel, cryogen-free, sensors that measure minute magnetic field variations (for details, see (6)). The majority of OPM-based MEG recordings (OPM-MEG) performed in humans have used alkali OPMs where either one (for single/dual-axis measurements (7)) or two (for tri-axial measurements (8)) photodetectors record the light intensity of a laser beam passing through a transparent cell containing ^87^Rb vapor heated to ∼150°C (Rb-OPMs) (7). Current implementations of Rb-OPMs lead to light (4.5–4.7 g) and small-size (1.2 × 1.7 × 2.6 cm^3^) OPMs with noise levels (i.e., <23 fT/rtHz in the 3–100 Hz frequency range) close to SQUIDs (i.e., 2–8 fT/rtHz), a bandwidth limited to below 130 Hz, a dynamic range limited to a few nT (<5 nT), single-to tri-axial magnetic field measurement (8), and heat dissipation power of ∼0.7 W per sensor (7). Tri-axial Rb-OPMs better differentiate environmental magnetic noise from neuromagnetic fields than single-axis Rb-OPMs, improving the efficiency of noise reduction techniques (9). Nevertheless, the radial magnetic component remains *a priori* the optimal choice to record the signal of interest as it is larger in amplitude and less affected by volume conduction than tangential components (10). Thanks to their reduced size, Rb-OPMs can be placed directly on or very close to the scalp while recording physiological brain activity (11-13) as well as epileptiform discharges in children (14-16) and adults (17, 18). In the case of pediatric epilepsy, the reduced brain-sensor distance afforded by on-scalp Rb-OPM-MEG led to higher IED amplitude and SNR compared with cryogenic MEG (14). The advent of Rb-OPM has thus ignited a revolution in the field of MEG and human neurosciences (7), and might—in time—become a reference method for the diagnostic evaluation of focal epilepsy (14, 19-22).

Despite these advantages, Rb-OPM-MEG suffers from some limitations that may limit their use for the study of human brain function (19). First, the high temperature of ^87^Rb vapor may constrain the number of sensors that can be applied on the scalp to ensure sufficient heat dissipation (23). It may also require increasing the scalp-sensor distance or placing thermal insulation to avoid discomfort. Second, atomic properties of ^87^Rb related to the spin-exchange rate intrinsically limit the recording bandwidth (6) (below 130 Hz at 150°C), precluding investigation of high frequency brain activity. They also limit the dynamic range of Rb-OPM sensors, imposing the need for on-board field nulling coils and strict magnetic shielding requirements, both passive (magnetic shielded room, MSR) and active (external coil systems (24, 25)). On-board field nulling is also required to avoid cross-axis projection errors (26).

An alternative to alkali OPM technology is offered by the optical pumping of He gas as sensitive element (He-OPMs) (27, 28). In the current prototype implementation used in this study, He-OPMs are heavier (40 g) and bigger (1.9 × 1.9 × 5 cm^3^) than Rb-OPMs, which therefore require a specific adaptable helmet to place them on the individuals’ head and limit the number of sensors that can be used. They have a higher noise level (i.e., <50 fT/rtHz over 1–1,500 Hz frequency range) but a larger dynamic range (beyond 200 nT) and bandwidth (0–2,000 Hz) that offers the opportunity to investigate high frequency brain oscillations, three axes of magnetic field measurement (although only two with noise <50 fT/rtHz), and they dissipate only ∼0.01 W per sensor as ^4^He needs no heating (29). He-OPMs have successfully recorded magnetic cardiac (30) and physiological brain (29, 31) activities. To the best of our knowledge, no study has so far demonstrated the ability of He-OPMs to record IEDs, nor has IED detection been compared between Rb- and He-OPM-MEG.

Both Rb- and He-OPMs benefit from three axes of magnetic field measurement at a single location that may prove particularly useful to increase spatial brain sampling and maximize the sensitivity of IEDs detection in epileptic patients. The number of OPMs that can be placed on the scalp is indeed restricted by technical (i.e., the heat dissipated by Rb-OPMs) or practical (i.e., the size/weight of He-OPMs) constraints.

This study therefore aims at demonstrating the practical interest of the three axes of measurements of Rb- and He-OPMs to maximize the detection of IEDS in one school-aged epileptic girl. It also aims at comparing the amplitude and SNR of IEDs simultaneously recorded with Rb- and He-OPMs. For that purpose, the patient underwent a multimodal electrophysiological recording comprising simultaneous scalp He-OPM-MEG (4 sensors) and Rb-OPM-MEG (4 sensors) alongside simultaneous scalp electroencephalography (EEG, 4 electrodes).

## Methods

### Case report

We studied a school-aged girl suffering from RFE, suffering from epilepsy for 4 years (Patient 5 in (14)). She underwent a right anterior temporal lobectomy leading to seizure-freedom (Engel class 1A) but IEDs remain very frequent with a maximal amplitude over C4-T4 electrodes (Figure 1). She underwent cryogenic MEG seven months prior to OPM-MEG recording, which showed right centrotemporal IEDs (Figure 1).

**Figure 1.**
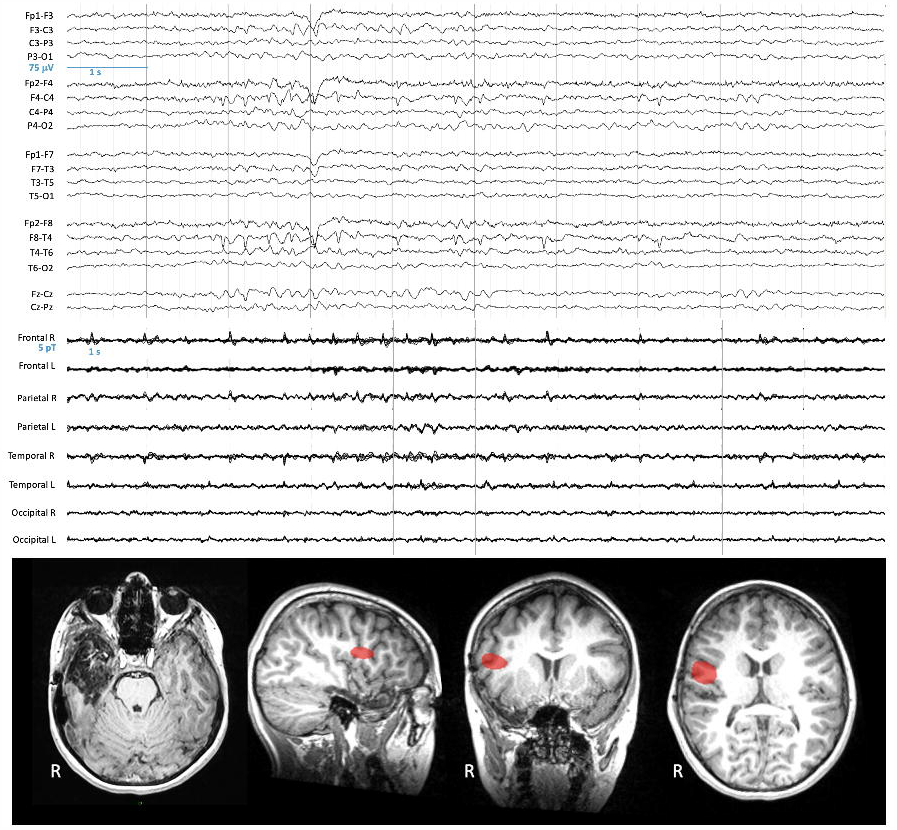
Clinical EEG data and previous cryogenic MEG data. **Top**. 10-second EEG signal (longitudinal bipolar montage) recorded during the clinical follow-up 7 months before the multimodal OPM acquisition, band-pass filtered at 0.53–70 Hz. **Middle**. Non-simultaneous 10 seconds of cryogenic MEG signals (magnetometers) recorded 7 months before the multimodal acquisition, band-pass filtered at 3–40 Hz. **Bottom**. Axial brain T1-weighted MRI illustrating the resection cavity after the resection of a right temporal dysembryoplastic neuroepithelial tumor (**Left**). Source localization of IEDs detected with cryogenic MEG signals (for methods, see ^17^) displayed on parasagittal (**Middle left**; right hemisphere), coronal (**Middle right**), and axial (**Right**) sections.

This study was approved by the institutional Ethics Committee (Hôpital Erasme, Reference: P2019/426). The patient and her legal representative gave written informed consent prior to the inclusion.

### Data acquisition

The patient underwent 40-minutes of multimodal electrophysiological recording based on four MEG-compatible EEG electrodes (silver Gold EEG disc electrodes, SPES Medica; placed at C4-T4-F8-Cz), five He-OPM prototypes (as described in (29)) and four Rb-OPMs (Gen-3.0, QuSpin Inc.; tri-axial mode, gain 2.7 V/nT; Figure 2). Still, poor EEG electrode impedances precluded qualitative analyses of EEG signals. OPMs were placed using an adaptable helmet (made from a photosensitive resin, designed for He-OPMs) placed on scalp and optimized for school-age children’s head size. Locations of the 89 3D printed sensor mounts (2 × 2 cm^2^) on the helmet were not based on conventional EEG montages (32) (Figure 2, left) but on specific locations optimizing the number of sensors. Four He-OPMs and four Rb-OPMs were placed in contact with the patient’s scalp at ∼1 cm from C4 or T4. One He-OPM was placed at the left centrotemporal region. Rb-OPMs were fixed to the sensor mounts using a layer of foam on the sensors’ sides as the sensor mount was too large (Figure 2, right), no layer of foam was added at the bottom of the sensor to maintain the same, virtual, scalp-to-sensor space compared with He-OPMs.

**Figure 2.**
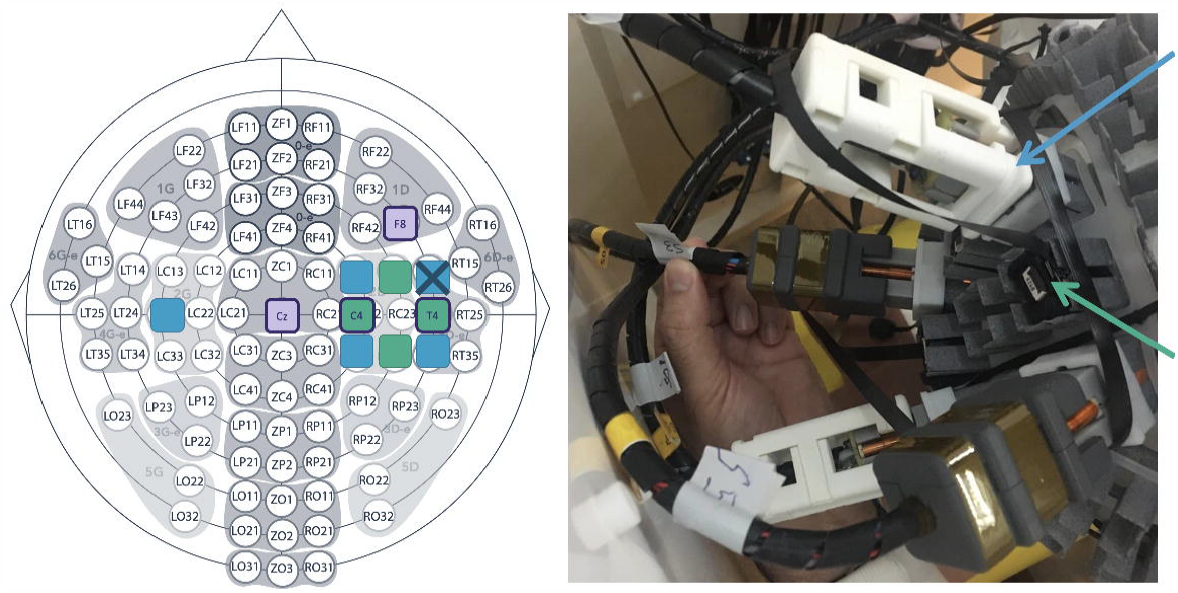
View of the multimodal EEG/He-OPM/Rb-OPM montage. **Left**. Map illustrating the placement of He-OPMs (blue), Rb-OPMs (green) and EEG electrodes (purple edges) with respect to a standard scalp EEG montage. The He-OPM marked with a cross was non-functional during the recording. **Right**. The OPMs were mounted on a dedicated helmet superimposed on scalp electrodes (not visible).

EEG signals were recorded using a commercially available EEG amplifier (Advance Neuro Technology, sampling rate 256 Hz, no band-pass filter). OPM signals were fed to distinct digital acquisition units (He-OPM, WeMEG Acquisition System SN001, sampling rate: 11,161 Hz, no band-pass filter; Rb-OPM, National Instruments DAC, sampling rate: 1200 Hz, no band-pass filter). A 1-Hz square-wave trigger signal was generated from the Rb-OPM acquisition system and sent to both He-OPM acquisition electronics and EEG amplifier to enable re-synchronization of the simultaneous recordings.

Recordings took place inside a compact MSR optimized for OPM recordings (Compact MuRoom, Cerca Magnetics Ltd, see (14)). Remnant magnetic field was reduced to 1–2 nT by combining degaussing and static background magnetic field compensation based on 22 field nulling coils placed within the MSR walls (cCoil, Cerca Magnetics Ltd, see (33)). The patient was free to move, comfortably sitting and watching a movie inside the MSR.

### Data preprocessing and analyses

Due to an unexpected technical problem, one of the four right-hemisphere He-OPM did not work properly and was excluded of subsequent analyses (Figure 2). The four other sensors were operating at a noise floor higher than usual (60–65 fT/rtHz). He-OPM data were first resampled at 1200 Hz with prior anti-aliasing low-pass filter (330 Hz), as were EEG data, and all acquisitions were then re-synchronized on the basis of the common trigger signal. He- and Rb-OPM data were then further band-pass filtered at 3–38 Hz (usual 3-40Hz band-pass filter dedicated to IED detection (14, 15) adapted to exclude an unprecedented 40Hz noise).

To isolate IED activity from background noise, independent component analysis (ICA) was performed on both Rb-OPM and He-OPM band-filtered signals separately (FastICA with nonlinearity *tanh*; see (34)). Components including IEDs were visually identified and the others (i.e., devoid of IEDs detectable by visual examination) were regressed out of OPM data. IED peaks were visually identified and counted in He- and Rb-OPM denoised data by three independent observers (O.F, F.W. and L.R.; Figure 3).

**Figure 3.**
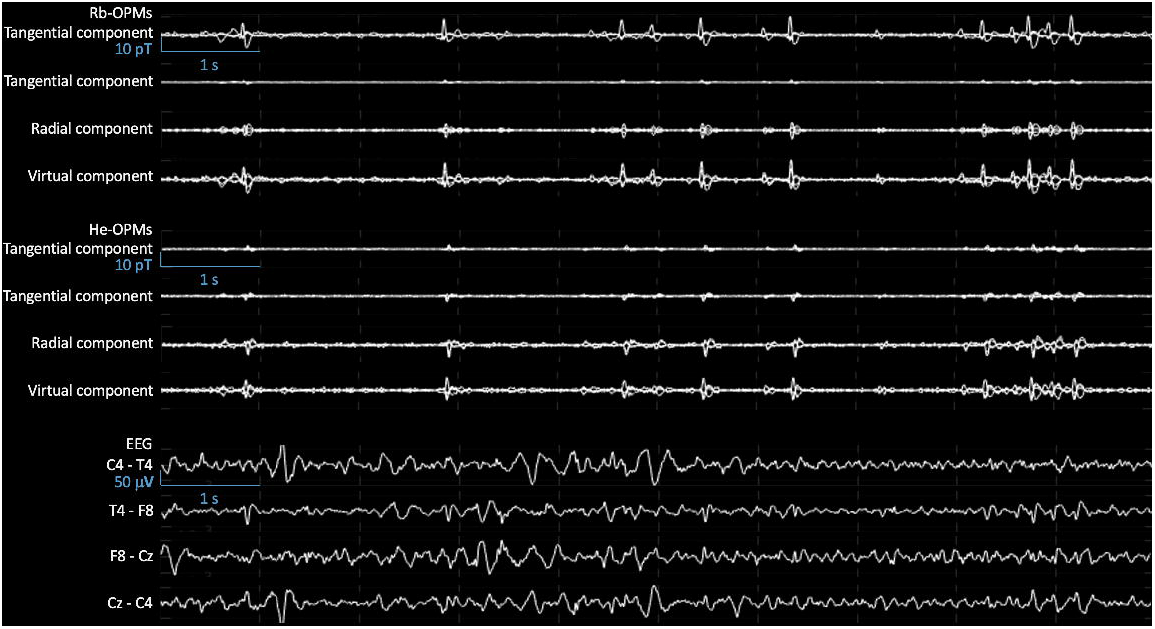
Sample of Rb-OPMs, He-OPMs, and EEG signals. Butterfly plots of 10-second signals of each tangential component, radial component, and virtual component from 4 Rb-OPMs (**Top**) and He-OPMs (**Middle**), after rejection of 11 independent components free of IEDs from the 12-channel raw data. Bipolar plots of 10-second simultaneous signals between each couple of EEG electrodes (placed according to the 10-20 montage) (**Bottom**). All signals were band-pass filtered at 3–38 Hz. This illustrates IEDs that can be detected simultaneously in both kinds of OPM.

Statistics were performed on a sub-selection of 102 IEDs simultaneously observed in artifact-free periods of Rb-OPM and He-OPM data, with IED amplitude being estimated at the peak of these IEDs. These analyses focused on the Rb- and He-OPM showing the highest IED amplitude (i.e., one Rb-OPM and one He-OPM out of 4, avoiding multiple comparisons). Given that the tri-axial measurements of two OPMs may vary just because of differences in their scalp location relative to IED source, steps were taken to assess and partly mitigate the effects of sensor positioning. First, the three magnetic components of each of the 102 IEDs considered for the analyses were compared using a one-way ANOVA and post-hoc *t* tests (significance at p < 0.05), so as to identify the most prominent field component at each sensor. Second, the best magnetic orientation was estimated at each sensor as the principal component of its tri-axial signals and then used to replace each tri-axial sensor by a virtual sensor projected along this orientation (Figure 3). Comparing amplitudes in these virtual sensors allowed for a principled comparison of two OPMs as it mitigates ambiguities related to sensor orientation. Peak amplitudes of each of the 102 IEDs in these virtual sensor data were then compared across modalities using two-sided paired *t* test (significance at p < 0.05).

To illustrate the denoising efficiency of ICA on IED recordings in both OPM modalities, the noise level of the ICA components that include IED activity was estimated from the background signal (i.e., artifact-free periods devoid of IEDs from 100 ms to 50 ms before each IED peak time) and compared statistically (two-sided paired *t* test at p < 0.05) with a similar estimate of noise level extracted from OPM data devoid from IED activity (i.e., obtained by regressing out ICA components that include IED activity). IED noise also enabled the comparison of peak IED SNR (i.e., the ratio of each peak IED amplitude to their corresponding background noise amplitude) across modalities (two-sided paired *t* test at p < 0.05).

## Results

To isolate IED activity from background noise, independent component analysis (ICA) was performed on both Rb-OPM and He-OPM band-filtered signals separately. Components including IEDs were visually identified and the others (i.e., devoid of IEDs detectable by visual examination) were regressed out of OPM data, leading to optimally-denoised versions with IED activity.

The ICA allowed to isolate IED activity in a single component, both with Rb- and He-OPMs. This yielded particularly clean signal traces for both OPM modalities (Figure 3). Three independent readers visually detected respectively 1372, 1287, and 1271 IEDs with Rb-OPMs and 1175, 1231, and 1221 IEDs with He-OPMs, most of them appeared simultaneously in both modalities on the corresponding ICA components (Figure 3).

The IED amplitude was significantly different across the three axes of the Rb- and He-OPMs (ANOVA, F_2,303_ =519, p=10^−98^, η^2^=79% for the selected He-OPM and F_2,303_=801, p=10^−121^, η^2^=84% for the selected Rb-OPM). It was significantly higher on one tangential axis for the Rb-OPM (first tangential component: 7.7pT +/- 0.18pT, radial component: 3.6pT +/- 0.08pT, second tangential component: 1.0pT +/- 0.02pT; post-hoc t-tests, |t_101_|=44, p=0, Cohen |d|=4.3) and on the radial axis for the He-OPM (radial component: 4.6pT +/- 0.15pT, tangential components: 1.5pT +/- 0.05pT and 2.0pT +/- 0.06pT; post-hoc t-tests, |t_101_|=31, p=0, Cohen |d|=3,1; Figure 4).

**Figure 4.**
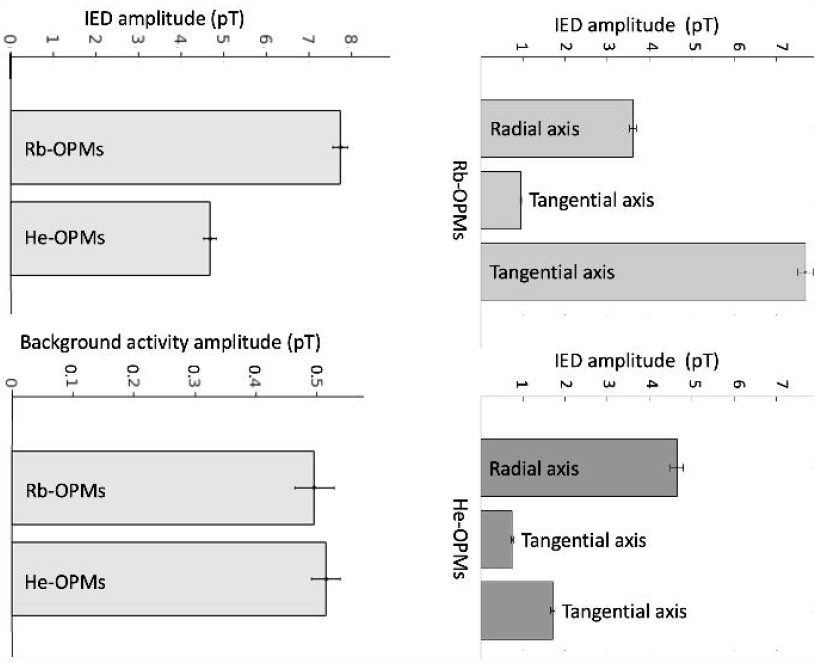
IED amplitude and background activity. **Top**. IED amplitude across three axes, after ICA preprocessing. Tri-axial Rb-OPM with the highest IED amplitude (light grey, **left**). Tri-axial He-OPM with the highest IED amplitude (dark grey, **right**). Amplitudes correspond to the single ICA component that contained IED activity. Bar plots show mean ± SD across a sample of 102 simultaneous IEDs. The IED amplitude was significantly higher on one tangential axis for the Rb-OPM and on the radial axis for the He-OPM, with respect to the relative position of each kind of OPM to the epileptogenic zone. These differences depending on the measurement axis can be explained by a more optimal position of Rb OPM than He OPM relative to the presumed irritative zone, as supported by the previous clinical EEG data. **Bottom**. IED amplitude, background activity after ICA preprocessing in the optimal magnetic orientation (virtual sensor). Comparison of each IED amplitude (**left**), background activity from 100 ms to 50 ms before each selected IED peak (**right**) between Rb-OPMs and He-OPMs. All amplitudes correspond to the virtual sensor signal built from the single ICA component that isolated IED activity. The two OPM modalities show similar background noise levels. The higher IED amplitude in Rb-OPM likely reflect difference in OPM positions with respect to the presumed irritative zone localization.

Given that ICA led to one-dimensional data with fixed IED magnetic orientation, we build virtual sensors that follow the time variations of magnetic field amplitude by suitable projection of each sensor three axes. These virtual sensors allowed to extract the IED amplitude (Rb-OPM: 7.8pT +/- 0.2pT, He-OPM: 4.7pT +/- 0.1pT; t-tests, |t_101_|=14, p=0, Cohen |d|=1,4; Figure 4) independently of sensor orientation and enabled their statistical comparison.

This revealed a higher IED amplitude and likewise a higher SNR (Rb-OPM: 21.3 +/- 1.4, He-OPM: 11.4 +/- 0.8; t-tests, |t_101_|=8, p=2×10^−11^, Cohen |d|=0,8) with the Rb-OPM sensor. Given the strong difference in IED field orientations at the Rb- and He-OPMs, the higher IED amplitude in the Rb-OPM probably reflected a better positioning relative to the IED neural source for the Rb-OPM compared to the He-OPM, rather than a difference in performance *per se*. In fact, background noise in the IED-specific ICA component was similar in the two modalities (Rb-OPM: 0.5pT +/- 0.03pT, He-OPM: 0.5pT +/- 0.02pT; t-tests, |t_101_|=1, p=0.26, Cohen |d|=0.1). This contrasted with the case of the other, IED-free, ICA components which exhibited higher background noise in the He-OPM (measured during background activity devoid of IEDs; Rb-OPM: 0.8pT +/- 0.1pT, He-OPM: 1.9pT +/- 0.2pT; t-tests, |t_22_|=4, p=2×10^−4^, Cohen |d|=0.9).

## Discussion

This case study demonstrates the ability of both He-OPM-MEG and Rb-OPM-MEG to record IEDs. Usage of ICA proved extremely efficient at denoising IED activity recorded with both Rb- and He-OPM-MEG, despite the low number of sensors, and led to similar remnant noise levels in both modalities despite the strong difference in intrinsic noise levels of the two types of OPMs (15 fT/rtHz for Rb-OPM vs. 65 fT/rtHz for He-OPM).

The lower IED amplitude and SNR reported for He-OPM-MEG is likely due to differences in sensor positioning rather than an intrinsically lower sensitivity. Indeed, Rb- and He-OPMs were placed at different scalp positions, with ∼2cm distance between OPMs (Figure 2). It is thus unsurprising that one modality (in this case, Rb-OPM) turned out advantaged with one sensor in a more optimal position than the other modality (He-OPM) to record the same focal brain activity. This was highlighted by the finding that the magnetic orientation was different in Rb-OPM (tangential orientation) and He-OPM (radial orientation). As radial magnetometers record brain activity originating around them and not just beneath (9), He-OPMs were likely placed on the sides of the IED source. Tangential field measurement allows the detection of a dipole just beneath the magnetometer (9), so the Rb-OPM with the highest IED amplitude was likely placed right above the IED source. This difference in OPM positioning further explains why IEDs detected by Rb-OPMs had higher amplitude and SNR than those recorded by He-OPMs. The difference in IED amplitude might also explain why slightly less IEDs (between 86 and 96%) were detected with He-OPMs than with Rb-OPMs, as a lower SNR complicates the unambiguous detection of low amplitude IEDs.

These data illustrate clearly, in a clinical setting, the benefits of tri-axial OPMs to maximize spatial brain sampling with a limited number of sensors (8). Apart from cost, the current size/weight of He-OPM prototypes and the heat dissipated by Rb-OPMs will indeed make it difficult to achieve full scalp coverage with a high number (>100) of OPMs packed close together. Thus, tri-axial OPMs will allow to reach a high number of recording channels with a reasonable number of OPMs placed on the whole scalp.

IEDs were successfully isolated within a single ICA component with both Rb-OPMs and He-OPMs. The efficiency of ICA in this context has been shown previously in MEG (34, 35). Our data provide the first demonstration of this efficiency in OPM-MEG, despite the small number of sensors used. This allowed to remove from the raw OPM signals a large part of the background activity (encompassing sensor noise, environmental noise, and physiologic brain signals unrelated to IEDs), to the point of leading to similar noise levels in the two types of OPMs. This procedure thus represents a promising approach to automatize and reduce the time allocated to visual IED detection (21, 36).

This study suffers from several limitations. First, it deserves confirmation in a larger population of epileptic patients with different forms of temporal and extra-temporal epilepsies. Second, it was limited by the low number of He- and Rb-OPMs that were placed at different locations to allow simultaneous recordings. This prevented proper spatial coverage of the IZ to enable IED source reconstruction, which would ultimately be the way to provide a comparison of Rb- and He-OPM-MEG free from the effect of relative positioning. Alternatives could be to swap sensor locations between He- and Rb-OPMs for a second recording or to perform consecutive recordings with He- and Rb-OPMs placed at similar locations. Still, these alternatives also suffer from some limitations such as increasing the recording duration that may impact patients’ cooperation (swapping and consecutive recordings) or that differences in IEDs amplitude/SNR may be due to variation in IEDs across time (consecutive recordings). Finally, the impact of simultaneously recording EEG electrodes on OPM SNR is also difficult to estimate.

Overall, this study highlights the added value of multi-axial OPMs when spatial sampling is limited. It also shows that Rb- and He-OPMs are both able to detect IEDs with similar noise levels on IED activity properly isolated with ICA, opening the door for the automatization of OPM-MEG data analyses in epileptic patients. Future clinical on-scalp OPM-MEG users should consider selecting the type of OPM to use depending on the balance between the benefits and disadvantages of each OPM technology.

## Abbreviations

EEG: Electroencephalogram
He-OPM: Helium-based optically pumped magnetometer
ICA: Independent component analysis
IED: Interictal epileptiform discharge
MEG: Magnetoencephalography
MSR: Magnetic shielded room
OPM: Optically pumped magnetometer
Rb-OPM: Rubidium-based optically pumped magnetometer
RFE: Refractory focal epilepsy
SNR: Signal-to-noise ratio
SQUID: Superconducting quantum interference device

## Acknowledgments

O.F. is supported by the Fonds pour la formation à la recherche dans l’industrie et l’agriculture (FRIA, Fonds de la Recherche Scientifique (FRS-FNRS), Brussels, Belgium). P.C. is supported by the Fonds Erasme (Convention « Alzheimer », Brussels, Belgium). X.D.T. is Clinical Researcher at the FRS-FNRS.

The OPM-MEG project at the Hôpital Universitaire de Bruxelles and Université libre de Bruxelles is financially supported by the Fonds Erasme (Projet de Recherche Clinique et Convention « Les Voies du Savoir 2 ») and by the FRS-FNRS (Crédit de Recherche: J.0043.20F, Crédit Équipement : U.N013.21F).

The MAGIC OPM MEG project at CEA Tech Lille, Amiens and Lille University Hospitals and Université de Picardie Jules Verne and Université de Lille was co-funded by European Union with the European Funding of Regional Development and the Region Hauts de France.

## Author contribution

O.F., E.L. and X.D.T. contributed to the design and conception of the study. O.F., P.C., M.M., C.S. and N.B. contributed to data acquisition. O.F., L.R., D.S., F.W., V.W. and X.D.T. contributed to data analyses. O.F., P.C., E.L., M.M., L.R., C.S., N.H., M.B., S.G., R.R., S.M., A.P.L., D.S., N.B., P.D., F.W., V.W. and X.D.T. contributed to drafting or reviewing the manuscript. E.L. and X.D.T. obtained funding.

## Data availability

Data are available upon reasonable request to the corresponding author and after approval of institutional (Hôpital universitaire de Bruxelles & Université libre de Bruxelles) authorities.

## References

1. Hämäläinen M, Hari R, Ilmoniemi RJ, Knuutila J, Lounasmaa OV. Magnetoencephalography theory, instrumentation, and applications to noninvasive studies of the working human brain. Reviews of Modern Physics. 1993;65.

2. Papanicolaou AC, Roberts TPL, Wheless JW. Fifty Years of Magnetoencephalography: Beginnings, Technical Advances, and Applications 2020.

3. Hari R, Puce A. MEG-EEG Primer 2017.

4. Vivekananda U. Redefining the role of Magnetoencephalography in refractory epilepsy. Seizure. 2020;83:70–5.

5. Boto E, Bowtell R, Kruger P, Fromhold TM, Morris PG, Meyer SS, et al. On the Potential of a New Generation of Magnetometers for MEG: A Beamformer Simulation Study. PLoS One. 2016;11(8):e0157655.

6. Tierney TM, Holmes N, Mellor S, Lopez JD, Roberts G, Hill RM, et al. Optically pumped magnetometers: From quantum origins to multi-channel magnetoencephalography. Neuroimage. 2019;199:598–608.

7. Boto E, Holmes N, Leggett J, Roberts G, Shah V, Meyer SS, et al. Moving magnetoencephalography towards real-world applications with a wearable system. Nature. 2018;555(7698):657–61.

8. Boto E, Shah V, Hill RM, Rhodes N, Osborne J, Doyle C, et al. Triaxial detection of the neuromagnetic field using optically-pumped magnetometry: feasibility and application in children. Neuroimage. 2022;252:119027.

9. Brookes MJ, Boto E, Rea M, Shah V, Osborne J, Holmes N, et al. Theoretical advantages of a triaxial optically pumped magnetometer magnetoencephalography system. Neuroimage. 2021;236:118025.

10. Iivanainen J, Stenroos M, Parkkonen L. Measuring MEG closer to the brain: Performance of on-scalp sensor arrays. Neuroimage. 2017;147:542–53.

11. Boto E, Seedat ZA, Holmes N, Leggett J, Hill RM, Roberts G, et al. Wearable neuroimaging: Combining and contrasting magnetoencephalography and electroencephalography. Neuroimage. 2019;201:116099.

12. Boto E, Hill RM, Rea M, Holmes N, Seedat ZA, Leggett J, et al. Measuring functional connectivity with wearable MEG. Neuroimage. 2021;230:117815.

13. Borna A, Carter TR, Colombo AP, Jau YY, McKay J, Weisend M, et al. Non-Invasive Functional-Brain-Imaging with an OPM-based Magnetoencephalography System. PLoS One. 2020;15(1):e0227684.

14. Feys O, Corvilain P, Aeby A, Sculier C, Holmes N, Brookes M, et al. On-Scalp Optically Pumped Magnetometers versus Cryogenic Magnetoencephalography for Diagnostic Evaluation of Epilepsy in School-aged Children. Radiology. 2022:212453.

15. Feys O, Corvilain P, Van Hecke A, Sculier C, Rikir E, Legros B, et al. Recording of Ictal Epileptic Activity Using on-Scalp Magnetoencephalography. Ann Neurol. 2023;93(2):419–21.

16. Feys O, Corvilain P, Bertels J, Sculier C, Holmes N, Brookes M, et al. On-scalp magnetoencephalography for the diagnostic evaluation of epilepsy during infancy. Clinical Neurophysiology. 2023.

17. Vivekananda U, Mellor S, Tierney TM, Holmes N, Boto E, Leggett J, et al. Optically pumped magnetoencephalography in epilepsy. Ann Clin Transl Neurol. 2020;7(3):397–401.

18. Hillebrand A, Holmes N, Sijsma N, O’Neill GC, Tierney TM, Liberton N, et al. Non-invasive measurements of ictal and interictal epileptiform activity using optically pumped magnetometers. Sci Rep. 2023;13(1):4623.

19. Brookes MJ, Leggett J, Rea M, Hill RM, Holmes N, Boto E, et al. Magnetoencephalography with optically pumped magnetometers (OPM-MEG): the next generation of functional neuroimaging. Trends Neurosci. 2022;45(8):621–34.

20. Pedersen M, Abbott DF, Jackson GD. Wearable OPM-MEG: A changing landscape for epilepsy. Epilepsia. 2022.

21. Feys O, De Tiege X. From cryogenic to on-scalp magnetoencephalography for the evaluation of paediatric epilepsy. Dev Med Child Neurol. 2023.

22. Feys O, Goldman S, Lolli V, Depondt C, Legros B, Gaspard N, et al. Diagnostic and therapeutic approaches in refractory insular epilepsy. Epilepsia. 2023;64(6):1409–23.

23. Hill RM, Boto E, Rea M, Holmes N, Leggett J, Coles LA, et al. Multi-channel whole-head OPM-MEG: Helmet design and a comparison with a conventional system. Neuroimage. 2020;219:116995.

24. Holmes N, Leggett J, Boto E, Roberts G, Hill RM, Tierney TM, et al. A bi-planar coil system for nulling background magnetic fields in scalp mounted magnetoencephalography. Neuroimage. 2018;181:760–74.

25. Iivanainen J, Zetter R, Gron M, Hakkarainen K, Parkkonen L. On-scalp MEG system utilizing an actively shielded array of optically-pumped magnetometers. Neuroimage. 2019;194:244–58.

26. Borna A, Iivanainen J, Carter TR, McKay J, Taulu S, Stephen J, et al. Cross-Axis projection error in optically pumped magnetometers and its implication for magnetoencephalography systems. Neuroimage. 2022;247:118818.

27. Beato F, Belorizky E, Labyt E, Prado ML, Palacios-Laloy A. Theory of a 4He parametric-resonance magnetometer based on atomic alignment. Physical Review. 2018.

28. Fourcault W, Romain R, Le Gal G, Bertrand F, Josselin V, Le Prado M, et al. Helium-4 magnetometers for room-temperature biomedical imaging: toward collective operation and photon-noise limited sensitivity. Opt Express. 2021;29(10):14467–75.

29. Labyt E, Corsi MC, Fourcault W, Palacios Laloy A, Bertrand F, Lenouvel F, et al. Magnetoencephalography With Optically Pumped (4)He Magnetometers at Ambient Temperature. IEEE Trans Med Imaging. 2019;38(1):90–8.

30. Morales S, Corsi MC, Fourcault W, Bertrand F, Cauffet G, Gobbo C, et al. Magnetocardiography measurements with (4)He vector optically pumped magnetometers at room temperature. Phys Med Biol. 2017;62(18):7267–79.

31. Gutteling TP, Bonnefond M, Clausner T, Daligault S, Romain R, Mitryukovskiy S, et al. A New Generation of OPM for High Dynamic and Large Bandwidth MEG: The 4He OPMs— First Applications in Healthy Volunteers. Sensors (Basel). 2023;23(2801).

32. Acharya JN, Acharya VJ. Overview of EEG Montages and Principles of Localization. J Clin Neurophysiol. 2019;36(5):325–9.

33. Holmes N, Rea M, Chalmers J, Leggett J, Edwards LJ, Nell P, et al. A lightweight magnetically shielded room with active shielding. Sci Rep. 2022;12(1):13561.

34. Vigario R, Sarela J, Jousmaki V, Hamalainen M, Oja E. Independent component approach to the analysis of EEG and MEG recordings. IEEE Trans Biomed Eng. 2000;47(5):589–93.

35. Malinowska U, Badier JM, Gavaret M, Bartolomei F, Chauvel P, Benar CG. Interictal networks in magnetoencephalography. Hum Brain Mapp. 2014;35(6):2789–805.

36. De Tiege X, Lundqvist D, Beniczky S, Seri S, Paetau R. Current clinical magnetoencephalography practice across Europe: Are we closer to use MEG as an established clinical tool? Seizure. 2017;50:53–9.

